# A New Approach to Identifying Elite Winter Sport Athletes’ Risk of Relative Energy Deficiency in Sport (REDs)

**DOI:** 10.1101/2024.09.27.24314508

**Authors:** Emily M. Smith, Kelly Drager, Erik M. Groves, Leigh Gabel, Steven K. Boyd, Lauren A. Burt

## Abstract

**Objectives:** Relative Energy Deficiency in Sport (REDs) is a syndrome resulting from problematic low energy availability (LEA). Low areal bone mineral density (aBMD) is a primary indicator of LEA, measured by dual X-ray absorptiometry (DXA). High-resolution peripheral quantitative computed tomography (HR-pQCT) is an advanced imaging device that provides measures of volumetric BMD (vBMD), bone microarchitecture, geometry, and strength. The objective of this study was to assess prevalence of REDs in elite winter sport athletes and to observe the associations in bone parameters using HR-pQCT in athletes identified as at-risk or not at-risk of REDs.

**Methods:** Participants included 101 elite athletes (24.1±4.4 SD years; 52% female). The REDs Clinical Assessment Tool (CAT2) was used to determine REDs risk. HR-pQCT scans of the non-dominant radius and left tibia were analyzed upon REDs risk grouping.

**Results:** Seventeen athletes (17%; 71% female) were at-risk based on the REDs CAT2. After covarying for lean mass, odds ratios (OR) suggested higher likelihood of REDs risk classification for athletes with low cortical thickness, cortical area, total vBMD, and bone strength.

**Conclusions:** Impaired total vBMD, bone strength and cortical bone parameters were approximately twice as likely (OR: 1.9-3.0) in athletes at-risk of REDs. Results agree with the consensus statement that HR-pQCT may identify impaired bone health in athletes at-risk of REDs. Future directions should use HR-pQCT to explore REDs risk longitudinally, utilizing bone change over time as this may provide greater insight. Using advanced imaging to explore REDs risk in a population of winter high-performance athletes is novel.

## Introduction

Effects of low energy availability (LEA) pose risks to an athlete’s health, wellness, and sports performance (1,2). LEA occurs when one’s energy expenditure outweighs their energy intake (3) and can be especially detrimental to bone health (4). Bone is a mechanosensitive tissue that requires adequate levels of physical stimulus and energy intake to induce positive adaptations (5,6) to maintain bone quality, including bone mineral density (BMD), strength, geometry, microarchitecture, and tissue mineralization. The relationship between bone health and LEA is well known (7), leading to the development of models to describe this relationship, including Relative Energy Deficiency in Sport (REDs) (8,9). In 2023, REDs was redefined as “a syndrome of impaired physiological and/or psychological functioning experienced by female and male athletes that is caused by exposure to problematic LEA” (9). A primary indicator of REDs risk is the emphasis on impaired bone health, specifically low BMD, as defined in the REDs Clinical Assessment Tool Version 2 (CAT2) (10).

Areal BMD (aBMD) is measured using dual X-ray absorptiometry (DXA); however, relying solely on DXA may be restrictive as the technology has limited ability to analyze bone strength and microarchitecture. High resolution peripheral quantitative computed tomography (HR-pQCT) is an advanced imaging technology that measures total, cortical and trabecular BMD, bone microarchitecture, geometry, and estimated strength using finite element analysis (FEA). Cortical bone microarchitecture and geometry may play a critical role in the development of bone stress injuries (BSIs) in athletes with menstrual dysfunction (12–14). Both compromised bone strength and cortical bone have been observed in amenorrheic athletes compared to both eumenorrheic athletes and non-athletes (16), thus associated decrements in strength and microarchitecture could be suggestive of REDs risk. Additionally, estimated strength derived by HR-pQCT and FEA has been shown to predict fracture risk independent of aBMD (11,15), which is crucial in preventative assessment for the longevity of an athlete’s career. The growing utility and emerging potential of HR-pQCT has led to the inclusion of this modality in the possible assessment of REDs risk, though further research is needed (17,18).

Bone quality measured by HR-pQCT in athletes at-risk of REDs is not well understood, especially among winter sport athletes. However, limited work suggests that bone health appears to be poorer in athletes at-risk of REDs compared with not at-risk athletes (19). Research in REDs risk primarily includes endurance athletes, as they tend to be more susceptible to REDs (20,21). The ability to use HR-pQCT in addition to DXA in identification of REDs risk is relatively unknown; however, assessing bone microarchitecture and strength (18) may provide greater detail and implications of impaired bone health.

This study aimed to determine the prevalence of winter sport athletes at-risk of REDs using the REDs CAT2. In addition, we explored the likelihood and association of HR-pQCT derived bone density, microarchitecture, geometry and estimated strength parameters observed in athletes at-risk versus not at-risk of REDs, based on the REDs CAT2. It was hypothesized that athletes considered at-risk of REDs would have inferior bone quality via HR-pQCT than those athletes not at-risk of REDs (22).

## Methods

### Participants

This cross-sectional study obtained ethics approval from the Conjoint Health Research and Ethics Board at the University of Calgary (REB19-1078). Data collection occurred between November 2019 and January 2024. Participants were recruited through collaborative efforts with the Canadian Sport Institute Alberta based on the following eligibility criteria: affiliation with the Canadian Sport Institute, competing at either elite/international (Tier 4) or world class (Tier 5) (23) level in a winter sport recognized as part of the Olympic Winter Games, at least 14 years old, and have not experienced pregnancy/lactation in the year before data collection. All participants provided informed consent prior to study participation. Individuals under 18 years were assessed and treated as mature minors and signed their own consent forms.

### Questionnaires

Participants completed electronic questionnaires capturing relevant information pertaining to REDs risk, including age at which athletes began training, fracture and BSI prevalence, and missed or modified training due to injury/fracture. For females, age of menses, cycle frequency/duration and use of contraception were collected. The REDs CAT2 Step 2 (10) was used to assess an athlete’s risk of REDs. Athletes within this cohort were classified into two rather than four groups: at-risk of REDs (yellow/“mild”, orange/“moderate”, or red/“severe”) or not at-risk of REDs (green/“none”). The REDs CAT2 captures information from blood-based data (i.e., testosterone, triiodothyronine (T3), and cholesterol) and mental health screening (i.e., disordered eating, anxiety and/or depression) though we did not have these data.

### Dual X-ray Absorptiometry (DXA)

DXA (GE Lunar iDXA, GE Healthcare) was used to measure aBMD (g/cm^2^) of the left hip (femoral neck (FN) and total hip (TH)), and lumbar spine (LS). In addition to aBMD, sex-, age-, and weight-matched Z-scores were calculated (iDXA, enCORE v18, GE Lunar). Total body DXA scans were conducted to obtain anthropometric measures including lean mass (i.e., skeletal muscle, organs, and connective tissue), fat free mass (i.e., bone, skeletal muscle, organs, and connective tissue), fat mass, and percent body fat. Precision errors of aBMD for DXA sites ranged from 0.51% to 1.14% (24).

### High Resolution Peripheral Quantitative Computed Tomography (HR-pQCT)

Participants completed unilateral HR-pQCT scans of the distal non-dominant radius and left tibia at a nominal isotropic resolution of 61 μm (XtremeCT II, Scanco Medical, Brüttisellen, Switzerland). If either site was unable to be scanned due to previous fracture or metal artifact, the contralateral limb was scanned. Scans captured 10 mm length totaling 168 slices and were located 9.5 mm and 22.5 mm proximal to the reference line for the radius and tibia, respectively (25).

Technicians performed and analyzed all scans using the standard manufacturer’s method (25) and subsequent fully automatic segmentation was applied prior to quantitative analysis (26). All scans were graded 1 to 5 for motion artifacts: a scan scoring ‘1’ had no motion, and a scan scoring ‘5’ was subject to severe blurring and discontinuities (27). Any scan with a motion score of ‘4’ or greater was excluded from analyses. Precision scores for HR-pQCT ranged from <2.4% for density to <3.3% for microarchitecture parameters, with the exception of cortical porosity (<13.7%) (28). Bone parameters including total (TtBMD; mg HA/cm^3^) and trabecular volumetric BMD (TbBMD; mg HA/cm^3^), trabecular number (TbN; mm^-1^), trabecular thickness (TbTh; mm), trabecular separation (TbSp; mm) and trabecular area (TbAr; mm^2^) were assessed using the standard morphologic analysis (25). Cortical parameters, including cortical volumetric BMD (CtBMD; mg HA/cm^3^), cortical thickness (CtTh; mm), cortical area (CtAr; mm^2^) and cortical porosity (CtPo; %), as well as total cross-sectional area (TtAr; mm^2^) were determined using an automated segmentation method (29,30). Image analysis was performed using Scanco Image Processing Language (version 6.6).

Estimated bone strength was determined via simulated failure load (FL) using finite element analysis (FEA) software (FAIM, version 9.0, Numerics88 Solutions, Calgary, Canada) (31). An axial compression test was simulated using a 1% compressive strain, Young’s modulus of 8748 MPa and a Poisson’s ratio of 0.3 (32). The primary estimate of bone strength was FL (kN) based on 2% of the elements exceeding 7000 microstrain (33). Precision errors are <2% (CV <1%) for estimates of stiffness and FL (34).

### Equity, Diversity and Inclusion

Recruitment of both male and female athletes in similar numbers was important to the application of the REDs CAT2 as males are often neglected in REDs research. Our team consists of academic researchers, exercise physiologists and a dietitian across all career stages with backgrounds in kinesiology, engineering and radiology.

### Patient and Public Involvement

Patients or the public were not involved in any aspect of this research.

### Statistical Analysis

Sample size was based on large effect sizes (0.9) previously observed between eumenorrheic and amenorrheic athletes and between athletic sporting groups using HR-pQCT (35,36). A minimum of 42 participants was required to achieve 80% power with a confidence level of 95% (α=0.05). Data were analyzed using R Statistical software (v.4.0.0). Continuous data were presented using mean and standard deviation and categorical data using numbers and percentages. Data were checked for normality using Shapiro–Wilk tests and visually checked with Q-Q plots and histograms. Demographic information was displayed for all athletes and split by sex, where two-sample t-tests were used to explore differences between at-risk and not at-risk groups. The primary outcome variables of this study included TtBMD and FL at both the radius and the tibia.

HR-pQCT and FEA results were compared with age- and sex-matched controls using freely available normative data (Normative^©^, https://www.normative.ca) developed and maintained by the Bone Imaging Laboratory at the University of Calgary and Z-scores were produced for each variable. Odds ratios (OR) utilized Z-scores for each HR-pQCT parameter and were used to determine likelihood and association between REDs risk and HR-pQCT parameters, where higher ORs represent a greater likelihood of observing a lower measure of each parameter (e.g., OR > 1: odds of observing low values of a parameter are greater in the at-risk group than in the not at-risk group). These models were adjusted for lean mass *a priori* as studies have found positive correlations between lean mass and aBMD (37). Due to sample size limitations, multiple variable logistic regression was performed, but restricted to a maximum of two variables, including the independent variable (e.g., TtBMD) and one covariate (i.e., lean mass).

## Results

### Demographics

One hundred and one elite winter sport athletes participated in this study (52% female) associated with the following winter sports: biathlon, cross country skiing, long track speed skating, figure skating and ski cross. Demographic and sport affiliation information are presented in Table 1. This cohort was 24.1±4.4 SD years old with an average training age of 13.2±6.3 SD years.

**Table 1.** Demographic Information for All Athletes and Proportion of Athletes by Sport.

| **Demographics** |  |  |  |
| --- | --- | --- | --- |
|  | **All** | **Male** | **Female** |
|  | **(N=101)** | **(n=48)** | **(n=53)** |
|  | Mean (SD) | Mean (SD) | Mean (SD) |
| Age (years) | 24.1 (4.4) | 24.0 (4.3) | 24.2 (4.6) |
| Height (cm) | 175.1 (8.6) | 180.8 (6.0) | 170.0 (7.3) |
| Weight (kg) | 72.0 (10.2) | 78.4 (7.9) | 66.2 (8.4) |
| Body mass index (kg/m<sup>2</sup>) | 23.4 (2.2) | 24.0 (2.3) | 22.8 (1.9) |
| Lean mass (kg) | 56.4 (9.3) | 64.3 (5.6) | 49.3 (5.3) |
| Fat mass (kg) | 12.7 (4.1) | 11.0 (3.0) | 14.2 (4.3) |
| Percent fat (%) | 17.7 (5.2) | 13.9 (2.8) | 21.1 (4.5) |
| Fat free mass (kg) | 59.4 (9.7) | 67.6 (5.9) | 52.0 (5.6) |
| Age of menarche (years) | NA | NA | 13.4 (1.4) |
| Training age (years) | 13.2 (6.3) | 12.8 (5.6) | 13.5 (6.9) |
| **Participants By Sport** | Number (%) | Number (%) | Number (%) |
| Biathlon | 14 (13.9%) | 5 (10.4%) | 9 (17.0%) |
| Cross country skiing | 23 (22.8%) | 12 (25.0%) | 11 (20.8%) |
| Figure skating | 10 (9.9%) | 4 (8.3%) | 6 (11.3%) |
| Long track speed skating | 33 (32.7%) | 17 (35.4%) | 16 (30.2%) |
| Luge | 6 (5.9%) | 2 (4.2%) | 4 (7.5%) |
| Ski cross | 15 (14.9%) | 8 (16.7%) | 7 (13.2%) |

Data are presented as mean (SD) for continuous variables and number (%) for discrete variables.

Athletes had DXA scans of the total body, hip and lumbar spine, and HR-pQCT scans of the radius and tibia; however, two athletes were without scans at the radius due to reported bilateral fractures. No scans were removed due to motion artifact.

### REDs Prevalence

Within this cohort, 17% of athletes (71% female) were considered at-risk of REDs and 83% (49% female) not at-risk by use of the REDs CAT2 risk. Of the 17% at-risk athletes, 88% (67% female) were considered yellow risk, 12% (100% female) were considered orange risk and zero were considered red risk. Due to sample size restrictions, yellow, orange and red risk were considered as “at-risk” in our study. Prevalence of REDs risk by sport type is displayed in Supplementary Table 1 and prevalence of each REDs CAT2 indicator for this cohort is displayed in Table 2. The majority of athletes at-risk of REDs were from biathlon (29%) and cross country skiing (26%). Information for athletes in each risk group and split by sex are displayed in Table 3.

**Table 2.**
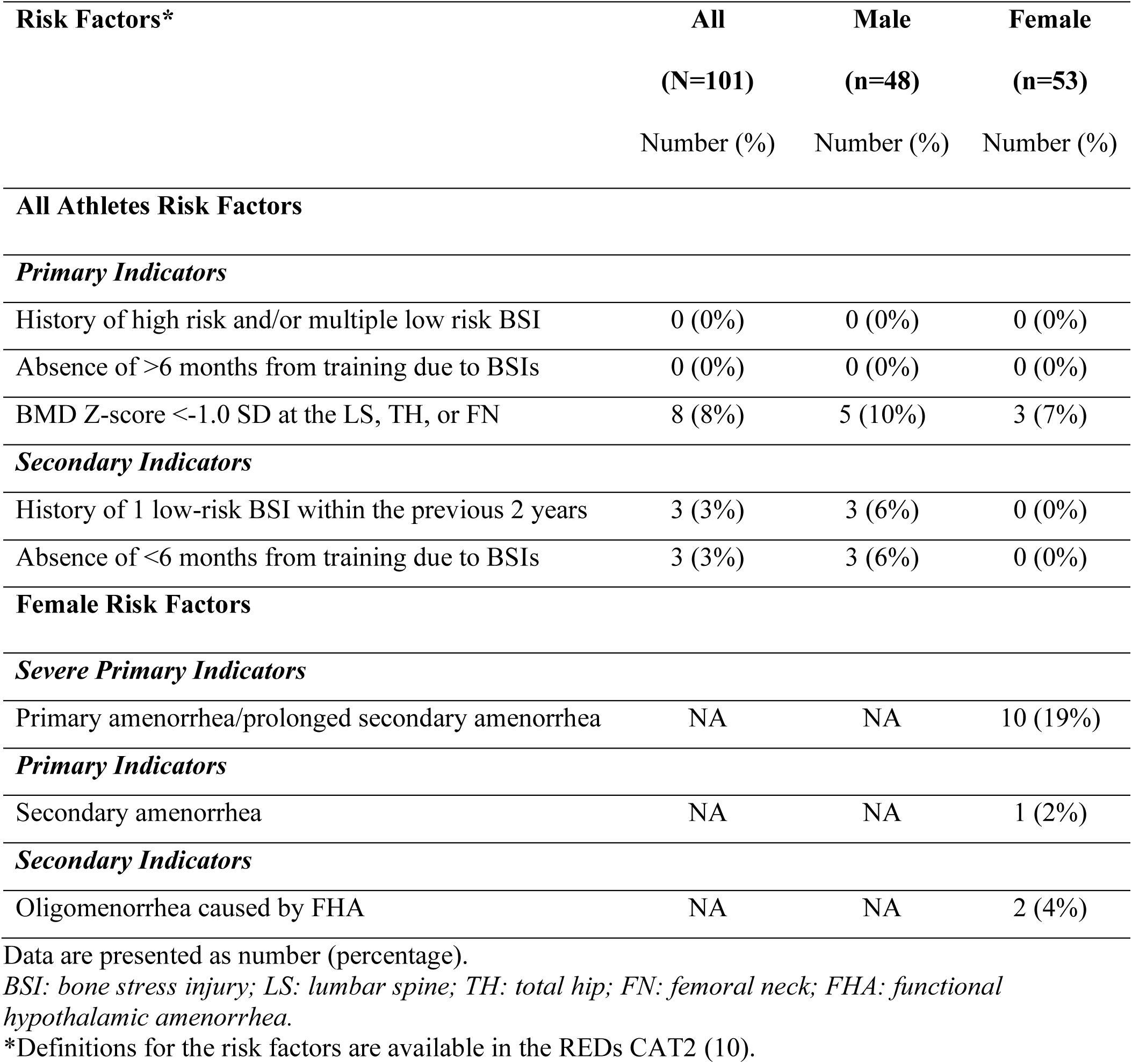
Prevalence of REDs CAT2 Indicators Used in Severity Classification Among Athletes Considered At-Risk of REDs.

| **Risk Factors\*** | **All**<br>**(N=101)**<br>**Number (%)** | **Male**<br>**(n=48)**<br>**Number (%)** | **Female**<br>**(n=53)**<br>**Number (%)** |
| --- | --- | --- | --- |
| **All Athletes Risk Factors** |  |  |  |
| *Primary Indicators* |  |  |  |
| History of high risk and/or multiple low risk BSI | 0 (0%) | 0 (0%) | 0 (0%) |
| Absence of >6 months from training due to BSIs | 0 (0%) | 0 (0%) | 0 (0%) |
| BMD Z-score <-1.0 SD at the LS, TH, or FN | 8 (8%) | 5 (10%) | 3 (7%) |
| *Secondary Indicators* |  |  |  |
| History of 1 low-risk BSI within the previous 2 years | 3 (3%) | 3 (6%) | 0 (0%) |
| Absence of <6 months from training due to BSIs | 3 (3%) | 3 (6%) | 0 (0%) |
| **Female Risk Factors** |  |  |  |
| *Severe Primary Indicators* |  |  |  |
| Primary amenorrhea/prolonged secondary amenorrhea | NA | NA | 10 (19%) |
| *Primary Indicators* |  |  |  |
| Secondary amenorrhea | NA | NA | 1 (2%) |
| *Secondary Indicators* |  |  |  |
| Oligomenorrhea caused by FHA | NA | NA | 2 (4%) |

Data are presented as number (percentage).
BSI: bone stress injury; LS: lumbar spine; TH: total hip; FN: femoral neck; FHA: functional hypothalamic amenorrhea.
\*Definitions for the risk factors are available in the REDs CAT2 (10).

**Table 3.** Demographics by REDs Risk Comparison for All Athletes and by Sex for At-Risk and Not At-Risk Groups.

|  | All Athletes |  |  | Male |  |  | Female |  |  |
| --- | --- | --- | --- | --- | --- | --- | --- | --- | --- |
|  | Not At-Risk<br>(n=84)<br>Mean<br>(SD) | At-Risk<br>(n=17)<br>Mean<br>(SD) | p-value | Not At-Risk<br>(n=43)<br>Mean<br>(SD) | At-Risk<br>(n=5)<br>Mean<br>(SD) | p-value | Not At-Risk<br>(n=41)<br>Mean<br>(SD) | At-Risk<br>(n=12)<br>Mean<br>(SD) | p-value |
| Age (years) | 23.9 (4.2) | 25.3 (5.8) | 0.346 | 24.1 (4.4) | 22.8 (3.0) | 0.339 | 23.6 (3.9) | 26.4 (6.4) | 0.195 |
| Height (cm) | 175.6 (8.4) | 172.7 (9.1) | 0.248 | 180.8 (6.1) | 180.3 (5.4) | 0.358 | 170.1 (6.8) | 169.6 (8.6) | 0.896 |
| Weight (kg) | **73.3 (10.0)** | **65.5 (7.5)** | **<0.001** | **79.2 (7.9)** | **71.7 (2.6)** | **<0.001** | 67.2 (8.4) | 62.9 (7.3) | 0.092 |
| BMI (kg/m<sup>2</sup>) | **23.7 (2.2)** | **21.9 (1.5)** | **<0.001** | **24.2 (2.3)** | **22.1 (1.8)** | **0.002** | **23.2 (2.0)** | **21.8 (1.4)** | **0.009** |
| Lean mass (kg) | **57.3 (9.5)** | **51.7 (6.6)** | **0.006** | **64.9 (5.6)** | **59.0 (1.9)** | **<0.001** | 49.4 (5.1) | 48.7 (5.3) | 0.712 |
| Fat mass (kg) | **13.0 (4.2)** | **11.1 (2.7)** | **0.027** | 11.1 (3.1) | 10.0 (1.7) | 0.810 | **14.9 (4.4)** | **11.6 (2.9)** | **0.005** |
| Percent fat (%) | 17.8 (5.5) | 17.0 (3.5) | 0.460 | 13.9 (2.9) | 13.8 (2.1) | 0.232 | **21.9 (4.5)** | **18.3 (3.2)** | **0.005** |
| FFM (kg) | **60.4 (9.9)** | **54.4 (6.8)** | **0.005** | **68.2 (5.9)** | **61.8 (1.9)** | **<0.001** | 52.2 (5.3) | 51.3 (5.6) | 0.681 |
| Menarche (years) | NA | NA | NA | NA | NA | NA | **12.9 (1.1)** | **15.1 (1.3)** | **<0.001** |
| Training age (years) | **12.2 (5.3)** | **17.6 (8.7)** | **0.017** | 11.2 (4.4) | 11.8 (5.4) | 0.784 | **13.4 (6.0)** | **20.0 (8.7)** | **0.022** |

*BMI: body mass index; FFM: fat free mass.*
Data are presented as mean (SD). P-values represent the difference between at-risk and not at-risk groups for all athletes, males and females.
\*Bolded values indicate statistical significance (p<0.05).

### Bone Quality Parameters

Adjusted ORs revealed that athletes with lower CtTh (OR radius: 2.1, p=0.021; tibia: 1.9, p=0.037) and CtAr (OR radius: 3.0, p=0.007; tibia: 2.7, p=0.006) were approximately two to three times more likely to be at-risk of REDs. Similarly, at the tibia, it was observed that athletes with lower TtBMD (OR tibia: 2.1, p=0.030) and FL (OR tibia: 2.2, p=0.033) were approximately twice as likely to be observed in the at-risk group. The OR results are displayed in Figure 1 and Supplementary Table 2.

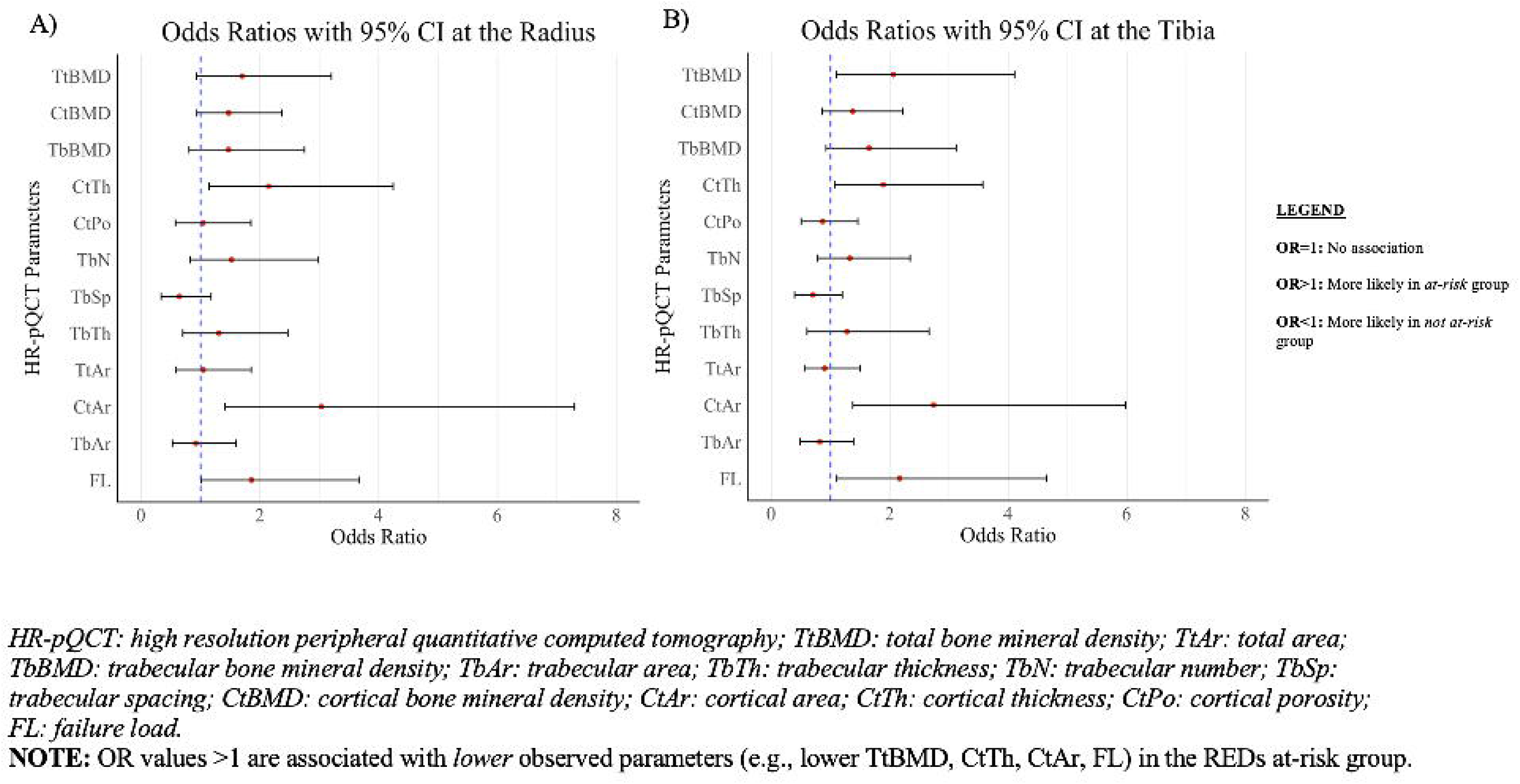

## Discussion

In our cohort, 17% of athletes were considered at-risk of REDs despite not being initially screened for risk (10). Previous research reports REDs prevalence between 15-80% depending on the sport (9,38), where leanness (or aesthetic) based sports and endurance sports tend to have higher prevalence due to the associated energy demands (20,39,40). This study included both endurance (i.e., cross country skiing, biathlon, and long track speed skating) and impact (i.e., figure skating, luge, ski cross) sports, and there was a higher percentage of athletes in the endurance-based sports considered at-risk, though the small sample size was underpowered to evaluate sport type differences. The inclusion of both endurance and impact sports may have contributed to the lower prevalence we report; however, we chose to include all sport types because studies involving winter sports are lacking. It is worth noting that athletes within this cohort were supported by a multi-disciplinary team at the Canadian Sport Institute Alberta, including performance dieticians and exercise physiologists, who are educated in REDs prevention strategies, possibly helping mitigate overall REDs risk.

Primary indicators for REDs include an aBMD Z-score <-1.0 SD at the lumbar spine, total hip or femoral neck, and a recent (i.e., within two years) history of high risk and/or multiple low risk bone stress injuries (BSI) (10). This cohort had a relatively small prevalence (8%) of athletes with low aBMD via DXA, zero athletes with a recent high-risk BSI and only 3% with a low-risk BSI. This differs from other studies which report 11-39% of athletes with low aBMD (22,41) and up to 20% of athletes sustaining at least one BSI per season (42). These disparities may be partially due to unique energy demands and loading patterns associated with winter sports compared with those usually studied (i.e., runners), and may also be partially attributed to the aforementioned support this cohort received.

Primary amenorrhea and prolonged secondary amenorrhea are defined in the REDs CAT2 as severe indicators of REDs for females (10). Previous studies report HR-pQCT differences in bone quality between athletes with and without menstrual dysfunction (13,16,36,43), as well as between athletes with and without BSIs (43). The development of BSIs in athletes with menstrual dysfunction has been linked to compromised cortical bone microarchitecture, geometry, and estimated strength (12–14), suggesting justification for HR-pQCT monitoring in athletes at-risk of REDs. These results are consistent with our findings whereby differences in HR-pQCT derived bone density (TtBMD), microarchitecture (CtTh), geometry (CtAr) and estimated bone strength (FL) were observed in athletes at-risk versus not at-risk of REDs. In our study, athletes with lower parameters were approximately twice as likely of being at-risk of REDs, supporting our hypothesis.

The current study did not observe differences between at-risk groups for trabecular bone; however, some studies have shown poorer quality of trabecular bone (i.e., higher separation, lower density, number and thickness) are observed in athletes with amenorrhea and/or LEA (16,36). The metabolic activity of trabecular bone makes it highly adaptive to mechanical loading, resulting in high potential for bone remodelling (44). The predominant gliding motion of these sports, in addition to the impact loading of some, may have masked or mitigated some of the negative effects of LEA on trabecular bone. It is also possible that changes in trabecular bone may be more affected by hormonal changes related to estrogen as seen in amenorrheic athletes (36,45,46), whereas in our cohort, only one at-risk athlete was classified based on secondary amenorrhea which might be most associated with an athlete’s current menstrual function (46). Notably, 70% (n=37) of the female athletes within this cohort were using contraceptives, thus the observance of secondary amenorrhea or oligomenorrhea in a natural cycle was not possible.

Future studies would benefit by exploring use of HR-pQCT in longitudinal analyses to observe REDs risk and associated changes in bone microarchitecture that may not otherwise be observed by DXA (17,47), as a negative change in aBMD Z-scores is also considered indicative of REDs risk (9,10). Additionally, further work should be done to include more impact or non- endurance sports for REDs CAT2 development (48).

### Clinical Implications

The release of the REDs CAT2 has provided clinicians with a means to assess athletes for REDs risk, despite REDs remaining a diagnosis by exclusion. This study contributes to the application required for the use of such a tool by applying it to a diverse group of athletes and sports. Based on these data, even with incomplete data for all indicators, the REDs CAT2 was used to identify athletes as being at-risk of REDs in a cohort that was not recruited for their REDs risk, nor was initial screening completed as per the REDs CAT2 Step 1 (10). These results suggest that even mild REDs risk may lead to impairments in bone quality, which may result in increased fracture risk. These data suggest an added benefit for the inclusion of HR-pQCT, in addition to DXA, for bone assessment in both research and clinical settings, to progress towards clinically relevant associations between REDs risk and bone health.

### Strengths/Limitations

This is the first study to apply the REDs CAT2 to winter sport athletes. This work contributes to the potential usage of HR-pQCT for studying various health outcomes, especially bone health, associated with REDs (9). However, the small number of athletes in the at-risk group and the incomplete panel of REDs CAT2 indicators limit the applicability and generalizability of these findings. Even with this tool, diagnosis is limited as all other potential clinical conditions must be systematically ruled out prior to confirming the presence of REDs (9,10), and without a single universal identifier, this makes risk classification inherently challenging. Additionally, this study compared bone quality by measure of HR-pQCT in a cohort where many had already been stratified as having low aBMD via DXA based on the REDs CAT2. While aBMD and HR-pQCT derived parameters are not linearly related in athletes (35), the reliance on aBMD to classify REDs risk may have confounded the results. However, not all athletes with low aBMD values had low HR-pQCT values. Future studies should examine the relationship between DXA and HR-pQCT skeletal outcomes and consider first stratifying athletes based on HR-pQCT parameters and subsequently evaluated for REDs risk. Finally, the cross-sectional nature of this study is limiting as bone metabolism may fluctuate throughout a competitive season (50) and longitudinal data may be more representative of athlete bone health.

## Conclusion

Athletes with lower cortical geometry and microarchitecture at both the radius and tibia, as well as lower total density and estimated strength at the tibia, were more likely to be identified as at-risk of REDs. Results from this study agree with the IOC consensus statement that HR-pQCT may identify impaired bone health in athletes at-risk of REDs. By using HR-pQCT prospectively, and screening all athletes for REDs risk, incidence of removal from sport due to extreme REDs risk classification may be reduced, increasing the longevity of the athlete.

## Statements and Declarations

The authors report no conflicts of interest in this work.

## Data Availability

All data produced in the present work are contained in the manuscript

## Acknowledgements

The authors would like to thank the athletes for their participation in this work. They would also like to thank the staff at the McCaig Institute for Bone and Joint Health, University of Calgary for their role in data scheduling and collection. They also wish to acknowledge the cooperative efforts of the Canadian Sport Institute Alberta in making this work possible.

## Contributions

Lauren Burt devised the study, assisted in data analysis and interpretation of results, and provided supervision throughout the study. They are the guarantor. Emily Smith contributed to athlete recruitment, data analysis, manuscript drafts, and creation of tables and figures. Kelly Drager helped with athlete recruitment, provided insight for study design and interpretation of results. Leigh Gabel helped provide insight for statistical analysis and interpretation of results. Erik Groves was involved in study design and interpretation of results. Steven Boyd assisted with data analysis and provided supervision throughout the study. All authors contributed to revision of this manuscript.

## Notes

### Competing Interest Statement

The authors have declared no competing interest.

### Funding Statement

This study did not receive any funding

### Author Declarations

This cross-sectional study obtained ethics approval from the Conjoint Health Research and Ethics Board at the University of Calgary (REB19-1078)

