## Supplementary Tables for "A New Approach to Identifying Elite Winter Sport Athletes’ Risk of Relative Energy Deficiency in Sport (REDs)"

**Supplementary Table 1.** REDs Risk by Sport Type.

| **Sport** | **At-Risk Athletes (n=17)** | **Total Athletes per Sport (N=101)** | **At-Risk % by Sport** |
|---|---|---|---|
| Biathlon | 4 | 14 | 29% |
| Cross Country Skiing | 6 | 23 | 26% |
| Figure Skating | 1 | 10 | 10% |
| Long Track Speed Skating | 5 | 33 | 15% |
| Luge | 1 | 6 | 17% |
| Ski Cross | 0 | 15 | 0% |

**Supplementary Table 2.** Odds Ratio Results for REDs Risk.

| | HR-pQCT Radius | | | HR-pQCT Tibia | | |
|---|---|---|---|---|---|---|
| **Predictor Variable** | **Odds Ratio** | **95% Confidence Interval** | **p-value** | **Odds Ratio** | **95% Confidence Interval** | **p-value** |
| TtBMD (mg HA/cm$^3$) | 1.7 | 0.9, 3.2 | 0.087 | **2.1** | 1.1, 4.1 | **0.030** |
| CtBMD (mg HA/cm$^3$) | 1.5 | 0.9, 2.4 | 0.098 | 1.4 | 0.9, 2.2 | 0.184 |
| TbBMD (mg HA/cm$^3$) | 1.5 | 0.8, 2.8 | 0.213 | 1.6 | 0.9, 3.1 | 0.103 |
| CtTh (mm) | **2.1** | 1.1, 4.2 | **0.021** | **1.9** | 1.1, 3.6 | **0.037** |
| CtPo (%) | 1.0 | 0.6, 1.8 | 0.889 | 0.9 | 0.5, 1.5 | 0.593 |
| TbN (1/mm) | 1.5 | 0.8, 3.0 | 0.195 | 1.3 | 0.8, 2.3 | 0.307 |
| TbSp (mm) | 0.6 | 0.3, 1.2 | 0.168 | 0.7 | 0.4, 1.2 | 0.209 |
| TbTh (mm) | 1.3 | 0.7, 2.5 | 0.397 | 1.3 | 0.6, 2.7 | 0.515 |
| TtAr (mm$^2$) | 1.0 | 0.6, 1.9 | 0.88 | 0.9 | 0.6, 1.5 | 0.672 |
| CtAr (mm$^2$) | **3.0** | 1.4, 7.3 | **0.007** | **2.7** | 1.4, 6.0 | **0.006** |
| TbAr (mm$^2$) | 0.9 | 0.5, 1.6 | 0.768 | 0.8 | 0.5, 1.4 | 0.449 |
| FL (kN) | 1.9 | 1.0, 3.7 | 0.056 | **2.2** | 1.1, 4.6 | **0.033** |

*Bolded values indicate statistical significance ($p<0.05$)

**NOTE:** OR values >1 are associated with *lower* observed parameters (e.g., lower TtBMD, CtTh, CtAr, FL) in the REDs at-risk group.
*HR-pQCT: high resolution peripheral quantitative computed tomography; TtBMD: total bone mineral density; TtAr: total area; TbBMD: trabecular bone mineral density; TbAr: trabecular area; TbTh: trabecular thickness; TbN: trabecular number; TbSp: trabecular spacing; CtBMD: cortical bone mineral density; CtAr: cortical area; CtTh: cortical thickness; CtPo: cortical porosity; FL: failure load.*
